# Predictive signs and symptoms of Bacterial Meningitis isolates in Northern Ghana

**DOI:** 10.1101/2022.05.27.22275694

**Authors:** Enoch Weikem Weyori, Nkrumah Bernard, Abass Abdul-Karim, John Bertson Eleeza, Braimah Baba Abubakari, Hilarius Asiwome Kosi Abiwu, Kuugbee Dogkotenge Eugene, Adadow Yidaana, Shamsu-Deen Ziblim, Benjamin Nuertey, Benjamin Asubam Weyori, Etowi Boye Yakubu, Stebleson Azure, Muktar Abdul-Muizz, Valentine Koyiri Cheba, Adatsi Richard Kojo

## Abstract

**Background:** Cerebrospinal meningitis (CSM) is a public health burden in Ghana that causes up to 10% mortality of the disease annually. About 20% of those who survive cerebrospinal meningitis suffer permanent sequelae. The study sought to understand the predictive signs and symptoms of bacterial meningitis implicated in its outcomes.

**Method:** Retrospective data from the Public Health Division, Ghana Health Service on bacterial meningitis from 2015 to 2019 used for this study. A pre-tested data extraction form was used to collect patients’ information from case-based forms kept at the Disease Control Unit from 2015 to 2019. Data were transcribed from the case-based forms into a pre-designed Microsoft Excel template. The data was cleaned and imported into SPSS version 26 for analysis.

**Results:** Between 2015-2019, a total of 2,446 CSM cases were reported. Out of these, 842 were confirmed. Among the confirmed cases, males constituted 55.3%. Children below 14 years of age were most affected (51.4%). The pathogens commonly responsible for bacterial meningitis were *Neisseria meningitidis* and *Streptococcus pneumoniae* with their respective strains especially *Nm W135, Nm X, Spn St. 1*, and *Spn St. 12F/12A/12B/44/4* being responsible for most of the confirmed cases. The most predictive sign and symptom for bacterial meningitis was fever (*X*^*2*^*=5*.*738*^*a*^; *p <0*.*05; AOR=1*.*303*). Identified signs that were associated but less likely to cause disease were abdominal pain (*X*^*2*^*=12*.*038*^*a*^; *p <0*.*05; AOR=0*.*597)*, neck stiffness (*X*^*2*^*=6*.*447*^*a*^; *p <0*.*05; AOR=0*.*788)*, altered consciousness (*X*^*2*^*=15*.*438*^*a*^; *p <0*.*05; AOR=0*.*673)*, and convulsion (*X*^*2*^*=15*.*084*^*a*^; *p <0*.*05; AOR=0*.*678)*.

**Conclusion:** The confirmed cases of bacterial meningitis from 2015 to 2019 had fever as the main predictive symptom. It is therefore noticed that abdominal pain, neck stiffness, altered consciousness and convulsion did not associate with CSM within the study period.

## Introduction

Meningitis stays one of the most pestilence inclined sickness influencing a huge extent of the total populace [1]. Bacterial meningitis is one of the most serious kinds of meningitis known to influence the focal sensory system in people. It is perceived as one of the best ten reasons for contamination related passing around the world, and around 30-half of the survivors support neurological sequelae [1]. Bacterial meningitis is likewise the commonest sort of meningitis overall and influences around 1.7 million individuals with roughly 170,000 passings yearly [1]. The most well-known specialists of bacterial meningitis are Haemophilus influenzae type b, Neisseria meningitidis, serogroups A, B, C, W135 and Y, and Streptococcus pneumoniae with various serotypes, for example, St1, St14, St19A and a lot more [1].

During the course of disease, microbes enter the subarachnoid space and the mind parenchyma through the circulation system or by direct intrusion through outer obstructions like the ear or sinuses [2]. The intrusion of the microorganisms sets off the arrival of cytokines, which prompt a fountain of provocative responses, bringing about serious irritation of the meninges encompassing the mind and the spinal line, prompting a perilous sickness [2]. Meningitis can likewise happen while liquid encompassing the meninges becomes contaminated [3]. The most well-known reasons for meningitis are viral and bacterial contaminations. Bacterial meningitis (traditionally, ‘pyogenic’ or polymorphonuclear meningitis) is a substantially more serious sickness than viral meningitis (traditionally ‘aseptic’ or lymphocytic) and on the off chance that untreated is very nearly 100% lethal [4]. Indeed, even with anti-infection treatment, bacterial meningitis stays a genuine reason for bleakness and mortality on the planet, and is a bacteriological crisis requiring dire finding and treatment [1].

Aside from bacterial meningitis, numerous different reasons for meningitis exist and incorporates viral meningitis, tuberculous meningitis (TBM) and other non-irresistible reasons for aseptic meningitis [5], [6]. Viral meningitis, which is generally less extreme than bacterial meningitis, is a consequence of meningeal contamination by different infections. An infection may just be recognized in half of the cases; the most well-known being enteroviruses [7]. Normal youth diseases, for example, chicken pox and measles have frequently been embroiled in viral meningitis [8]. TBM is brought about by Mycobacterium tuberculosis and is an extremely serious type of spread tuberculosis. Like intense bacterial meningitis, TBM additionally brings about high paces of neurological intricacies and frequently deep rooted sequelae [1]. Without appropriate treatment the death pace of TBM can be exceptionally high [9]. Aseptic meningitis is a term saved for the meningitis for which starting clinical assessment and routine research facility tests (counting Gram staining and CSF culture) neglect to uncover a clear reason [10]. The etiology of aseptic meningitis frequently incorporates viral, contagious or TBM. Non-irresistible reasons for aseptic meningitis like malignancies with cerebrum metastasis or a few prescriptions; remarkably sulphamethoxazole and nonsteroidal mitigating drugs, have additionally been recognized [11], [12].

A key element that adds to this high grimness is the deficient comprehension of the pathogenesis of the illness bringing about it being one of the main sources of mortality on the planet. Overcomers of bacterial meningitis can experience the ill effects of genuine neurological confusions, for example, deafness, visual deficiency, mental and scholarly disability which frequently endure over the lifetime of the person [13]. Regardless, the African meningitis belt (AMB) is a bunch of 26 nations extending between Senegal toward the west to Ethiopia in the east. These nations are known for having a high yearly rate of bacterial meningitis [14]. It is assessed that, frequency rates during pandemics have reached as high as 100-1000 cases for every 100,000 which are remarkably high rates for an obtrusive bacterial infection. The case casualty rate goes from 6.6 to 10.0% and extremely durable sequelae including hearing misfortune and engine hindrance influencing more than 30% of survivors, if untreated [15].

In Ghana, Neisseria meningitidis, Streptococcus pneumonia and Haemophilus influenzae stay the most common species found around the meningitis belt during episode circumstances and the non-pandemic circumstances too [16]. Bacterial Meningitis is a general wellbeing trouble in Ghana and can cause mortality up to 10% of the impacted people yearly. Around 20% of the people who endure the infection experience super durable sequelae [17]. The review pointed toward giving quantitative evaluation of the prescient signs and symptoms of bacterial meningitis in Ghana and to give results that is summed up over the entire Ghanaian populace.

## Method

### Study Design

This was a retrospective, cross-sectional study conducted using a consolidated database from the Disease Control and Surveillance Unit and the Zonal Public Health Laboratory, Tamale. The independent variables were signs and symptoms of the patient

### Study Setting

The study data covered all the districts and regions in Ghana. The study was carried out at the Zonal Public Health Laboratory, Tamale (ZPHL). ZPHL is a reference laboratory for bacterial meningitis in Ghana and West Africa. It serves as the reference public health laboratory for the northern zone of Ghana.

### Study Population

All patients who presented with signs and symptoms suggestive of bacterial meningitis per the Ghana Health Service case definition for bacterial meningitis were included in the study. All samples collected across the country are brought to the ZPHL for testing and or confirmation.

### Case Definition

#### Clinical case definition

An illness with sudden onset of fever (>38.5°C rectal or >38.0°C axillary) and one or more of the following: neck stiffness, altered consciousness, another meningeal sign or petechial or purpureal rash (GHS, 2019). In patients less than one (1) year, suspect meningitis when fever accompanied by bulging fontanelle (GHS, 2019).

#### Laboratory criteria for diagnosis

Positive CSF antigen detection or Positive culture at district or facility level and confirmed by PCR test within 72 hours of disease presentation (GHS, 2019).

#### Case classification

Suspected case is that which meets the clinical case definition, probable case is a suspected case as defined above and turbid CSF (with or without positive Gram stain) or ongoing epidemic and epidemiological link to a confirmed case, and confirmed case is a suspected or probable case with laboratory confirmation.

### Sample Size determination

No sample size was determined as all suspected cases of bacterial meningitis were included in the study from 2015-2019.

### Data Collection, Entry and Analysis

Data was extracted from the case reporting forms unto a pre-designed Microsoft excel template. The data was cleaned twice and exported to *SPSS Version 25*, for analysis. Descriptive analysis was performed and presented in graphs and tables. A chi-square analysis was performed for associated signs/symptoms of bacterial meningitis whiles binary logistics regression model was adopted to determine the clinical signs and symptoms that are predictive of a person likely to be tested positive for bacterial meningitis using the five years retrospective data. The dependent variable remained to be the test outcomes for the Polymerase Chain Reaction (PCR) (*Positive and Negative*).

### Inclusion Criteria

All patients that fulfilled the case definition criteria for bacterial meningitis were included to the study as collated by the Disease Control and Surveillance Unit and the ZPHL.

### Exclusion Criteria

All patients with inadequately filled case investigation forms, cases that were not having samples accompanying the case investigation forms and cases that had no culture and PCR results.

### Ethical Consideration

Ethical clearance for the study was sought from the Kwame Nkrumah University of Science and Technology Ethical Review Board with a reference number of CHRPE/AP/469/20. Permission was also sort from the Regional Health Administration and the Zonal Public Health Laboratory with reference number of GHS/ZPHL/0014/20.

## Findings and Results

A total of 2446 suspected cases were documented within the study period of which males were predominant (52.7%). The upper west and northern regions recorded the highest cases within the study period (40.6%) (Table 1). Majority of the study population were within 15-49 (40.1%) years (Table 1).

**Table 1:** Demographic Characteristics of the cases recorded over the five years period.

| **Indicator** | **Case Count** | **Percentage** |
| --- | --- | --- |
| **Sex** |  |  |
| Male | 1288 | 52.7 |
| Female | 1158 | 47.3 |
| **Total** | **2446** | **100** |
| **Regions** |  |  |
| Northern | 993 | 40.6 |
| Upper East | 460 | 18.8 |
| Upper West | 993 | 40.6 |
| **Total** | **2446** | **100** |
| **Age Grouping** |  |  |
| Under 5 | 442 | 18.1 |
| 5 – 14 | 673 | 27.5 |
| 15 - 44 | 981 | 40.1 |
| 45 - 59 | 182 | 7.4 |
| 60 + | 168 | 6.9 |
| **Total** | **2446** | **100** |

### Regional Distribution of Suspected Bacterial Meningitis cases

The northern and upper west regions recorded the highest and lowest suspected cases of meningitis in 2015 and 2016 respectively (Figure 1). Between 2017-2019 however, the upper west region consistently recorded more suspected cases than the other two regions (Figure 1)

**Figure 1:**
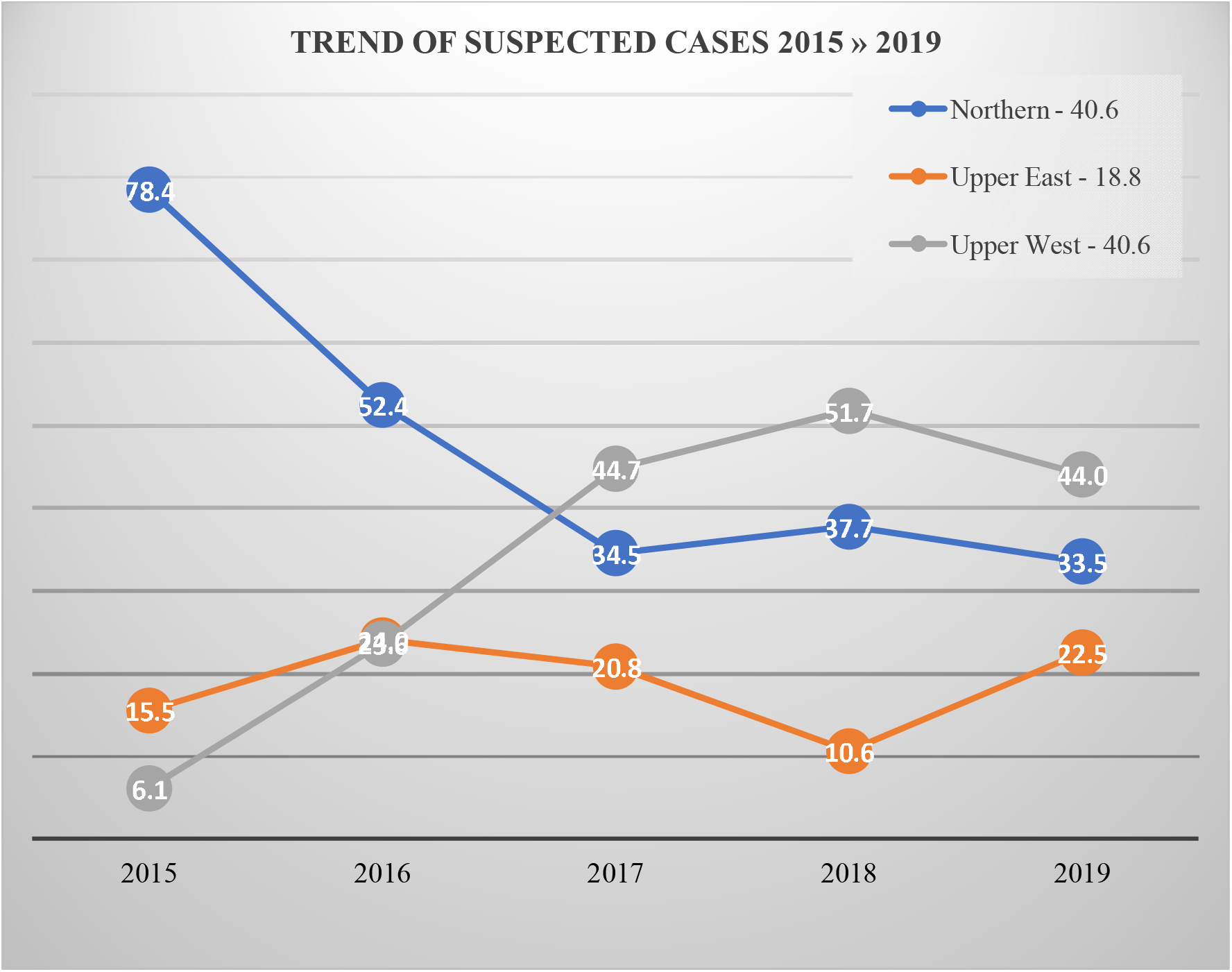
Regional counts of suspected cases across the years under review

### Trend of Confirmed Bacterial Meningitis cases from 2015 - 2019

The total case positivity within the study period was 34.4% (842/2446) with 2017 having the highest positivity rate of 35.7% (301/2446) and 2015 the lowest positivity rate of 6.9% (58/2446) (Table 2). Northern region recorded more cases than the other two regions across the study period (Table 2).

**Table 2:** Shows the distribution of cases across the years (Rt-PCR Outcomes)

| *PCR Outcomes* | *2015* | *2016* | *2017* | *2018* | *2019* | *Total* |
| --- | --- | --- | --- | --- | --- | --- |

| *Outcomes of suspected cases* |  |  |  |  |  |  |
| --- | --- | --- | --- | --- | --- | --- |
| *Negative* | 155 | 95 | 488 | 385 | 481 | 1604 |
| *% of N* | 72.8 | 38.6 | 61.9 | 69.1 | 75.0 | 65.6 |
| *Positive* | 58 | 151 | 301 | 172 | 160 | 842 |
| *% of N* | 27.2 | 61.4 | 38.1 | 30.9 | 25.0 | 34.4 |
| ***Total*** | **213** | **246** | **789** | **557** | **641** | **2446** |
| *Regional distribution of confirmed cases* |  |  |  |  |  |  |
| *Northern* | 49 | 88 | 142 | 95 | 53 | **427** |
| *% of NOR* | 84.5 | 58.3 | 47.2 | 55.2 | 33.1 | **50.7** |
| *Upper East* | 7 | 27 | 61 | 31 | 51 | **177** |
| *% of UER* | 12.1 | 17.9 | 20.3 | 18.0 | 31.9 | **21.0** |
| *Upper West* | 2 | 36 | 98 | 46 | 56 | **238** |
| *% of UWR* | 3.4 | 23.8 | 32.6 | 26.7 | 35.0 | **28.3** |
| ***Total*** | **58** | **151** | **301** | **172** | **160** | **842** |
| *Age Category of confirmed cases* |  |  |  |  |  |  |
| *0 – 14* | 31 | 76 | 168 | 89 | 67 | 433 |
| *% of 0-14* | 53.4 | 50.3 | 55.8 | 51.7 | 41.9 | 51.4 |
| *15 – 44* | 21 | 51 | 97 | 66 | 71 | **306** |
| *% of 15-44* | 36.2 | 33.8 | 32.2 | 38.4 | 44.4 | **36.3** |
| *45 – 59* | 4 | 8 | 14 | 10 | 10 | **46** |
| *% of 45-59* | 6.9 | 5.3 | 4.7 | 5.7 | 6.3 | **5.5** |
| *60+* | 2 | 4 | 22 | 7 | 12 | **45** |
| *% of 60+* | 3.4 | 2.6 | 7.3 | 4.1 | 7.5 | **5.3** |
| ***Total*** | **58** | **151** | **301** | **172** | **160** | **842** |

Overall, the 0-14 years age category had the highest positivity rate across the study period with the highest rates recorded in 2017 (Table 2)

### Signs and symptoms associated with bacterial meningitis

Fever (72.9%), neck stiffness (69.3%), and headache (56.6%) were the most reported signs and symptoms. Bulging fontanelle (5.6%), breathing difficulties (6.5%), abdominal pains (8.1%), back pain (7.4%) and waist pains (5.3%) were also reported (Table 3).

**Table 3:** Shows the Signs and Symptoms presented by patients across the five years period

| Signs and Symptoms | Present sign (N=2446) |  | Presence of BM (N=842) |  | Test of Association |  |
| --- | --- | --- | --- | --- | --- | --- |
| | frequency | % | frequency | % | $\chi^2$ | Sig. |
| Fever | 1783 | 72.9 | 611 | 72.6 | 5.738<sup>a</sup> | .018 |
| Neck stiffness | 1695 | 69.3 | 569 | 67.6 | 6.447<sup>a</sup> | .011 |
| Headache | 1384 | 56.6 | 506 | 60.1 | 6.450<sup>a</sup> | .012 |
| Bulging Fontanelle | 137 | 5.6 | 40 | 4.8 | 1.756<sup>a</sup> | .196 |
| Convulsion | 538 | 22 | 223 | 26.5 | 15.084<sup>a</sup> | .000 |
| Altered Consciousness | 526 | 21.5 | 219 | 26.0 | 15.438<sup>a</sup> | .000 |
| Breathing Difficulty | 158 | 6.5 | 53 | 6.3 | 0.058<sup>a</sup> | .863 |
| Abdominal Pains | 197 | 8.1 | 90 | 10.7 | 12.038<sup>a</sup> | .001 |
| Diarrhoea | 300 | 12.3 | 114 | 13.5 | 1.938<sup>a</sup> | .173 |
| Dizziness | 264 | 10.8 | 84 | 10.0 | 0.890<sup>a</sup> | .373 |
| Vomiting | 327 | 13.4 | 125 | 14.8 | 2.418<sup>a</sup> | .133 |
| Waist Pains | 130 | 5.3 | 53 | 6.3 | 2.449<sup>a</sup> | .129 |
| Loss of Appetite | 247 | 10.1 | 73 | 8.7 | 2.885<sup>a</sup> | .091 |
| Back Pain | 181 | 7.4 | 78 | 9.3 | 6.509<sup>a</sup> | .012 |
| Cough | 254 | 10.4 | 90 | 10.7 | .128<sup>a</sup> | .728 |
| Kennings Signs | 272 | 11.1 | 113 | 13.4 | 6.874<sup>a</sup> | .010 |
| Photophobia | 268 | 11 | 83 | 9.9 | 1.590<sup>a</sup> | .220 |

Bacterial meningitis was confirmed for majority of the Fever (72.6%), neck stiffness (67.6%), and headache (60.1%) signs and symptoms (Table 3).

### Signs and symptoms predictive of bacterial meningitis

Fever (*p=0*.*041*), neck stiffness (*p=0*.*026*), convulsion (*p=0*.*007*), abdominal pain (*p=0*.*043*), and altered consciousness (*p=0*.*000*) were associated with bacterial meningitis whiles others such as bulging fontanelle (*p=0*.*350*), headache (*p=0*.*098*), breathing difficulty (*p=0*.*950*) and diarrhoea (*p=0*.*388*) were not predictive of bacterial meningitis (Table 4).

**Table 4:** Shows the Binary Logistics Model summary for COR and AOR in the equation

| Variables in the Equation | B | S.E. | Wald | df | Sig. | COR(B) | AOR(B) | 95% C.I. for EXP(B) |  |
| --- | --- | --- | --- | --- | --- | --- | --- | --- | --- |
|  |  |  |  |  |  |  |  | Lower | Upper |
| Fever | .193 | .099 | 3.792 | 1 | .041 | 1.213 | 1.303 | 1.009 | 1.474 |
| Neck Stiffness | -.218 | .098 | 4.928 | 1 | .026 | .804 | .788 | .663 | .975 |
| Headache | -.152 | .092 | 2.739 | 1 | .098 | .859 | .803 | .717 | 1.028 |
| Bulging Fontanelle | .198 | .212 | .873 | 1 | .350 | 1.219 | 1.291 | .804 | 1.848 |
| Convulsion | -.291 | .108 | 7.297 | 1 | .007 | .748 | .678 | .605 | .923 |
| Altered Consciousness | -.416 | .109 | 14.618 | 1 | .000 | .660 | .673 | .533 | .817 |
| Breathing Difficulty | -.012 | .188 | .004 | 1 | .950 | .988 | 1.043 | .684 | 1.427 |
| Abdominal Pains | -.339 | .168 | 4.091 | 1 | .043 | .713 | .597 | .513 | .990 |
| Diarrhoea | -.122 | .141 | .747 | 1 | .388 | .885 | .838 | .672 | 1.167 |
| Dizziness | .170 | .154 | 1.211 | 1 | .271 | 1.185 | 1.141 | .876 | 1.604 |
| Vomiting | .023 | .134 | .029 | 1 | .865 | 1.023 | .826 | .787 | 1.331 |
| Waist Pains | -.333 | .195 | 2.925 | 1 | .087 | .717 | .751 | .489 | 1.050 |
| Loss of Appetite | .170 | .162 | 1.103 | 1 | .294 | 1.185 | 1.282 | .863 | 1.627 |
| Back Pain | -.199 | .167 | 1.417 | 1 | .234 | .819 | .672 | .590 | 1.137 |
| Cough | .032 | .155 | .041 | 1 | .839 | 1.032 | .952 | .761 | 1.399 |
| Kenigns Signs | -.186 | .143 | 1.682 | 1 | .195 | .830 | .710 | .627 | 1.100 |
| Photophobia | .254 | .158 | 2.573 | 1 | .109 | 1.289 | 1.192 | .945 | 1.759 |
| Constant | .302 | .383 | .621 | 1 | .431 | 1.352 | .525 |  |  |

Source: Field data, 2020.

## Discussion

The burden of bacterial meningitis is disproportionately distributed by place, time, and age but nearly equally distributed by sex. Within the study period, a total of two thousand, four hundred and forty-six (2446) cases were recorded and tested by both culture and Real Time Polymerase Chain Reaction (Rt-PCR) methods for pathogens responsible for bacterial meningitis. The trends of the suspected cases increased over the years with 2015 recording 8.6% of the cases to 25.9% in 2019. The results further revealed that 52.7% of the suspected cases were males. In addition, the Rt-PCR testing confirmed 34.4% of the suspected cases as bacterial meningitis. It further showed that, 55.3% of the confirmed bacterial meningitis cases were males whiles 44.7% were females. It was worth noting that 2017 had the highest total case positivity rates of 30.7%, while 2015 had the least total case positivity rate of 8.6%. This finding clearly shows that males in all instances have higher rates compared to females and this could be as a result of population dynamics of the respective regions under the study where males to females ratio is estimated at approximately 93% (Ghana Statistical Service, 2014). Also, it could be as a result of stronger immune system in women than men against viruses and bacteria infections (Chiaroni-Clarke et al., 2016). This finding is discordant with a similar study conducted by Kwambana-Adams et al., (2016) on pneumococcal meningitis outbreak and its associated factors in six districts of Brong Ahafo region, Ghana (Kwambana-Adams et al., 2016). The researchers documented that 55.9% of the confirmed cases were females compared to 44.1% males. This disparity could be due to the study period within which both studies were conducted. Whilst our study stan a five-year period (2015-2019), the other was done within a year (2015-2016).

Geographical distribution of confirmed cases across the five-year period denoted a general increase in confirmed cases of bacterial meningitis. The study revealed that over the five-year period, the Northern and Upper West regions reported the highest number of suspected cases (993; 40.6%) respectively whiles Upper East had the least suspected cases with 466 (18.8%). Further analysis reveals that, out of these suspected cases across the three regions, the northern region had 50.7% of confirmed cases of bacterial meningitis compared to the Upper East region which recorded a positivity rate of 21.0%. The UWR also recorded significant positivity rate of 28.3%. These findings are consistent with a study done by Codjoe & Nabies, (2014). The authors shared that the suspected and confirmed case of bacterial meningitis were highest in Northern region and Upper West regions within the meningitis belt in Ghana (Codjoe, S. N. A., & Nabie, 2014).

Meningitis cases was higher among younger age groups and adults below 44 years. This could be due to the increased likelihood of these groups of people participating in activities within overcrowded places such as schools, markets and other workplaces as well as type of settlement. The finding is in line with Amadu et. al., 2019 who had similar outcomes and trends in the demographic features of the cases (Amidu et al., 2019). The most suspected and confirmed cases of the bacterial meningitis in these regions remains among the 15 – 44 years age group with a total of 981 cases (40.1%). The confirmation of suspected cases by Rt-PCR denoted that child within the 0-14 age bracket had a total of 52.4% positivity rate compared to adults within the 15-60 age group that had a positivity rate of 41.8% (Amidu et al., 2019). These outcomes might be as a result of the vulnerability of children to infectious diseases of which bacterial meningitis is not an exception. This supports the argument that children younger than 15 years of age accounts for majority of all infections across the world (Conen, A., Walti, L. N., Merlo, A., Fluckiger, U., Battegay, M. & Trampuz, 2008; Falade, A. G., Lagunju, I. A., Bakare, R. A., Odekanmi, A. A. & Adegbola, 2009; Franco-Paredes, C., Lammoglia, L., Hernandez, I. & SantosPreciado, 2008; Saez-Llorens, X. & McCracken, 2003; World Health Organization, 2005b). Our finding is also comsistent with that of Nyarko (2016) who identified that 77.3% (761/980) of the confirmed meningitis cases were among children below the ages of 30 years in the Upper West region (Nyarko et al., 2016).

In 2016 a total of 61.4% of the suspected cases were positive for bacterial meningitis compared to the other years. The number of confirmed positive cases over the period denoted an increased pattern from 2015 with 6.9% of the total positives to 35.7% in 2018. This pattern dropped sharply in 2019 to 19.0% indicating a significant decline in cases over the one-year period. This patterns and trends seen over the period is in congruent with a study conducted on the US Centers for Disease Control and Prevention’s (CDC) surveillance data on bacterial meningitis from 1998 to 2003, where there was a significant reduction in the incidence of cases of bacterial meningitis cases (Thigpen, M. C., Whitney, C. G., Messonnier, N. E., Zell, E. R., Lynfield, R., Hadler, J. L. et al., 2011).

The common signs and symptoms, fever (72.9%), neck stiffness (69.3%), and headache (56.6%) were associated with bacterial meningitis. Other significant signs and symptoms identified were convulsion (22.0%), altered consciousness (21.5%), abdominal pain (8.1%) and back pain (7.4%). Binary logistics regression of the signs and symptoms revealed that fever (*p*=0.041; AOR=1.303), neck stiffness (*p*=0.026; AOR=0.788), convulsion (*p*=0.007; AOR=0.678), altered consciousness (*p*=0.00; AOR=0.673), abdominal pain (*p*=0.043; AOR=0.597) and back pain (*p*=AOR=0.672), were associated with bacterial meningitis. This aligns with an earlier publication by the CDC in 2012 that fever, headache, stiff neck, nausea, vomiting, photophobia, altered mental status remain as the major signs and symptoms of bacterial meningitis (CDC, 2012).

This study found that fever is likely to be caused by bacterial meningitis than any other signs and symptoms presented by cases (AOR=1.303; C.I =1.009-1.474). The other associated signs and symptoms were negatively associated with bacterial meningitis. This clearly indicates that fever is the only strong factor associated with bacterial meningitis within the study period and location. This outcome could be as a result of the high temperatures experience by the three northern zones of Ghana which exposes the people to the extreme heat.

## Conclusions and Recommendation

Bacterial meningitis continues to be an important cause of morbidity and mortality throughout the world, with differential risk among gender, age and geographic location. There is an increase in the rates of the disease pathogen over the period of the study. Children aged 0 -14 years, males and northern region are the most affected. Fever, neck stiffness, convulsion, altered consciousness abdominal pains, backpain and kennings signs are general signs associated with bacterial meningitis with fever been the main predictive sign and symptom for bacterial meningitis.

We recommended that peripheral health facilities should be keen in the identification of predictive signs and symptoms with particular attention to fever. This study could also inform Ghana Health Service in the review of protocols stipulated for managing suspicion levels of clinicians in managing cases of bacterial meningitis.

## Data Availability

Yes, all data are available and confidential under oath with the research team and availability will be on request

## Acknowledgement

The authors wish to thank the leadership of Ghana Health Service, Public Health Division and the Disease Surveillance Unit for their support and guidance. We also want to thank the leadership of the ZPHL add the rest for the immense support given to us through the period of data collection and processing.

## Authors’ contributions

The authors of the research contributed in diverse areas with specific roles and responsible as denoted;

^1*^Enoch Weikem Weyori, contributed in document management and write up

^2^Nkrumah Bernard, contributed in document review

^3^Abass Abdul-Karim contributed in document review

^4^John Bertson Eleeza contributed in document review

^5^Braimah Baba Abubakari contributed in document review

^6^Hilarius Asiwome Kosi Abiwu contributed in document review

^7^Kuugbee Dogkotenge Eugene contributed in document review

^8^Adadow Yidaana contributed in document review

^9^Shamsu-Deen Ziblim contributed in document review

^10^Benjamin Nuertey contributed in literature review

^11^Benjamin Asubam Weyori contributed in data management

^12^Etowi Boye Yakubu contributed in data management and analysis

^13^Stebleson Azure contributed in laboratory testing and results management

^14^Valentine Koyiri Cheba contributed in laboratory testing and results management

^15^Adatsi Richard Kojo contributed in laboratory testing and results management

